# Comparison of a prototype SARS-CoV-2 lateral flow immunoassay with the BinaxNOW™ COVID-19 Antigen CARD

**DOI:** 10.1101/2022.09.16.22279736

**Authors:** Haydon J. Hill, Timsy Uppal, Derrick Hau, Sujata G. Pandit, Jose Arias-Umana, Abigail J. Foster, Andrew Gorzalski, Kathryn J. Pflughoeft, Amanda R. Burnham-Marusich, Dana E. Reed, Marcellene A. Gates-Hollingsworth, Lynette Gumbleton, Subhash C. Verma, David P. AuCoin

## Abstract

**Background:** Robust diagnostics, capable of detecting multiple variant of SARS-CoV-2 are necessary to mitigate the COVID-19 pandemic. In this study we directly compare the diagnostic capabilities of an LFI engineered with monoclonal antibodies (mAbs) originating from SARS-CoV-2 NP immunizations to the Abbott BinaxNOW™ COVID-19 Antigen CARD.

**Methods:** Here we established a library of 18 mAbs specific to SARS-CoV-2 NP and used two of these mAbs (1CV7 and 1CV14) to generate a prototype antigen-detection lateral flow immunoassay (LFI). Samples consisting of remnant RT-PCR positive patient nasopharyngeal swabs preserved in viral transport media (VTM) were tested on the 1CV7/1CV14 LFI and the commercially available BinaxNOW™ test. Assays were allowed to resolve and results were recorded by two observers.

**Findings:** A total of 98 remnant SARS-CoV-2 positive patient specimens were tested on both the 1CV7/1CV14 LFI and the BinaxNOW™ test. The 1CV7/1CV14 LFI detected 71 of the total 98 specimens, while the BinaxNOW™ test detected 52 of the 98 specimens. Additionally, the 1CV7/1CV14 LFI consistently detected samples with higher RT-PCR cycle threshold values than the BinaxNOW™ test.

**Interpretation:** The 1CV7/1CV14 LFI outperformed the BinaxNOW™ test in the detection of BA.2, BA.2.12.1, and BA.5 Omicron sub-variants when testing remnant RT-PCR positive patient nasopharyngeal swabs diluted in viral transport media. BA.1 and BA.4 detection was comparable. The data suggest that mAbs derived from SARS-CoV-2 NP can aid in a more sensitive diagnostic immunoassay for COVID-19.

**Funding:** The study was funded by the University of Nevada, Reno’s Research and Innovation Office, DxDiscovery, Inc. internal funds, and through AuCoin Laboratory internal funds.

**Research in Context:** *Evidence before this study:* Since the onset of the pandemic, rapid antigen tests have proven themselves to be an accessible, accurate diagnostic platform. The widespread distribution of these tests has aided in curbing the COVID-19 pandemic. Data has shown that the tests manufactured at the beginning of the pandemic, utilizing monoclonal antibodies (mAbs) isolated from severe acute respiratory syndrome coronavirus (SARS-CoV), are less sensitive at detecting severe acute respiratory syndrome coronavirus 2 (SARS-CoV-2) Omicron and Omicron subvariants. The reduced sensitivity can lead to diagnostic escape, and possible surges in COVID-19 caseloads

*Added value of this study:* In this study, a total of 98 remnant RT-PCR confirmed SARS-CoV-2 positive clinical specimens were tested on both a prototype rapid antigen test in the form of a lateral flow immunoassay (LFI) (referred to as the 1CV7/1CV14 LFI) and the available Abbott BinaxNOW™ COVID-19 Antigen CARD. The 1CV7/1CV14 LFI detected markedly more specimens (71 of 98) specimens than the BinaxNOW™ test (52 of the 98).

*Implications of all the available evidence:* This research suggests that that the use of mAbs isolated from immunizations with protein from SARS-CoV-2 may result in a diagnostic assay that is more sensitive in detection of SARS-CoV-2 Omicron subvariants, in comparison to the existing BinaxNOW™ COVID-19 Antigen CARD.

## Introduction

Severe acute respiratory syndrome coronavirus-2 (SARS-CoV-2) is the causative agent of COVID-19.(1) As of September 2^nd^, 2022, the virus has caused an estimated 600 million infections and over 6.4 million deaths.(2) To contain the pandemic, the FDA has approved vaccines, therapeutics, and diagnostics in record time.(3–5) While this has served to slow hospitalizations and deaths, viral transmission has not been halted. Data has shown that social distancing and quarantining of infected individuals is one of the most effective ways to combat the spread of SARS-CoV-2.(6) In order to effectively quarantine, broad availability of accurate and rapid diagnostic tests is critical.

SARS-CoV-2 is a positive-sense single-stranded RNA virus that encodes for multiple genes that are expressed following infection. Various viral targets are used for diagnosing SARS-CoV-2 infections. Nucleic acid amplification tests (NAATs), such as polymerase chain reaction (PCR), primarily detect viral genomic RNA that encodes the spike and nucleocapsid proteins. (7) Antigen tests, such as lateral flow immunoassays (LFIs), primarily detect viral proteins such as the nucleocapsid protein (NP).

The gold standard diagnostic for SARS-CoV-2 infection are NAATs.(8) However, RNA viruses have a rapid mutation rate, and SARS-CoV-2 is no exception. B.1.617.2 (Delta) and B.1.1.529 (BA.1, Omicron) have developed many mutations, which can result in immune evasion.(9–11) The cumulative mutations acquired by the Delta and Omicron variants resulted in 9- and 36-amino acid replacements, respectively, in the spike protein alone, in reference to the original WA1 strain.(12,13) Because NAATs use nucleic acid primers to recognize and amplify their targets, inclusion of one or more base changes in the targeted nucleotide sequence is potentially enough to inhibit the reaction. The proliferative mutations acquired by the Omicron variant were enough to reduce clinical sensitivity with certain NAATs, leading to diagnostic escape. As a result, the Food and Drug Administration (FDA) ultimately recommended against using two NAATs for SARS-CoV-2 detection.(14–16)

In general, lateral flow immunoassays (LFIs) are inexpensive and user-friendly diagnostic assays that can be used at the point of care (POC).(17,18) Currently, LFIs are widely used to detect SARS-CoV-2 NP in patient samples, which potentially leads to a rapid COVID-19 diagnosis. In contrast to the spike protein, the NP is relatively conserved from variant to variant, containing only 3- and 6-amino acid changes for Delta and Omicron, respectively, in reference to the original WA1 strain.(12,13) The recent cluster of Omicron subvariants, BA.2, BA.2.12.1, and BA.5 all contain the same 7-amino acid replacements in the NP, 6 of which are conserved with the BA.1 variant.(19,20) Omicron subvariant BA.4 has yet another amino acid change, resulting in 8 replacements in the NP.(21) As a consequence, detection of the NP instead of the spike protein should result in an assay that is more resilient to diagnostic escape. In fact, every rapid detection test approved under Emergency Use Authorization by the FDA since 2020 is still being recommended for SARS-CoV-2 detection.(14) The pandemic presented an unparalleled need for rapid testing. To accommodate this, companies developed rapid tests utilizing mAbs previously isolated from severe acute respiratory syndrome coronavirus (SARS-CoV) NP immunizations.(22,23) These rapid tests performed well, as the homology between SARS-CoV NP and SARS-CoV-2 NP from the WA1 strain is 90%, and the use of mAbs generated to SARS-CoV NP allowed for the rapid production of assays to detect SARS-CoV-2 NP in patient samples.(24) However, with more SARS-CoV-2 variants appearing and the relatively slow yet continuous accumulation of mutations in the NP, the need for mAbs and diagnostic tests developed specifically for SARS-CoV-2 NP may be a necessity to maintain high diagnostic efficacy.

Presented is a prototype LFI purposely designed for the detection of SARS-CoV-2 NP in clinical specimens. A library of 18 mAbs generated through immunization with the SARS-CoV-2 NP was developed. The reactivities of these mAbs were determined by enzyme-linked immunosorbent assays (ELISA). LFIs were constructed and screened using mAb pairs to develop an assay with high analytical sensitivity and minimal background signal. Following the selection of the top mAb pairs and assay optimization, the LFI prototype was tested using cultured live viruses isolated from patient nasopharyngeal swabs to determine analytical sensitivity across variants. The LFI was subsequently tested using clinical specimens preserved in viral transport media (VTM), and was directly compared to Abbott BinaxNOW™ COVID-19 Antigen CARD. In general, the 1CV7/1CV14 LFI achieved a higher level of detection of Omicron and Omicron subvariants, compared to the BinaxNOW™ test.

## Methods

### Ethics statement

Laboratory animal work was approved by the University of Nevada, Reno Institutional Animal Care and Use Committee (Protocol #20-06-1026). All animal work was overseen by the Office of Laboratory Animal Medicine, which complies with the National Institutes of Health Office of Laboratory Animal Welfare policies (Assurance #A3500-01).

Deidentified human specimens (nasopharyngeal swabs) were used for the remnant clinical specimen testing. All the experiments were done in accordance with guidelines of the University of Nevada, Reno. The University of Nevada, Reno Institutional Review Board (IRB) reviewed this project and determined this study to be EXEMPT FROM IRB REVIEW according to federal regulations and University policy. The Environmental and Biological Safety committee of the University of Nevada, Reno, approved the methods and techniques used in this study.

### Monoclonal antibody production

Female 8-week-old BALB/c mice (Charles River Laboratories, Inc., Frederick, MA, RRID: IMSR_CRL:028) were immunized intraperitoneally with recombinant SARS-CoV-2 (WA1 strain) NP (The Native Antigen Company, United Kingdom) using Freund’s complete adjuvant (MilliporeSigma, Billerica, MA). Following the initial priming injection, subsequent immunizations were performed through intraperitoneal injections of SARS-CoV-2 (WA1 strain) NP (RayBiotech, Peachtree Corners, GA) mixed with Freund’s incomplete adjuvant (MilliporeSigma). Serum samples were collected through retro-orbital bleeds, and titers determined through indirect ELISA with SARS-CoV-2 (WA1 strain) NP (Thermo Fisher Scientific, Waltham, MA) immobilized to the plate. Hybridoma fusions were performed using a standard protocol.(25) Monoclonal antibodies (mAbs) were purified from the cell supernatant utilizing standard protein A affinity chromatography.

### Indirect ELISA

Microtiter 96-well flat-bottom medium binding plates (Greiner Bio-One, Austria) were coated with recombinant SARS-CoV-2 (WA1 strain) NP (Thermo Fisher Scientific) diluted in 1X DPBS (Corning, Corning, NY) overnight at room temperature. Plates were then washed three times with 1X PBS containing 0.05% Tween 20 (PBS-T). Plates were then blocked for 90 minutes at 37 °C in 1X PBS 0.5% non-fat milk and 0.1% Tween 20 (blocking buffer). Primary antibodies (mouse sera, hybridoma supernatant, or purified mAb) were diluted in blocking buffer and subsequent serially across the plate. Plates incubated in primary solution for 90 minutes at room temperature and were then washed three times with blocking buffer. Secondary antibody of either horseradish peroxidase-labeled polyclonal goat anti-mouse IgG antibodies (SouthernBiotech, Birmingham, AL, RRID: AB_2619742) or IgG subclass-specific polyclonal goat anti-mouse antibodies (SouthernBiotech) diluted in blocking were incubated in the wells at room temperature for 60 minutes. Plates were washed three times with PBS-T. Tetramethylbenzidine (TMB) 2-component peroxidase substrate (SeraCare, Milford, MA) was added to plates and allowed to react for 30 minutes at room temperature, then stopped using 1M H_3_PO_4_. Plates were read at OD 450.

### Lateral flow immunoassay (LFI) prototype development

The 18 purified NP-reactive mAbs were sprayed individually on CN95 nitrocellulose membranes (Sartorious, Gottingen, Germany) using a BioDotXYZ3060 dispense system (BioDot, Irvine, CA) at a concentration of 1 mg/mL for the test line. Goat anti-mouse IgG (SouthernBiotech, RRID:AB_2794121) was sprayed at 1 mg/mL for the control line. Sprayed nitrocellulose was attached to the LFI backing card (DCN, Carlsbad, CA) along with a CSFP203000 wicking pad (MilliporeSigma, Billerica, MA). 17 of the 18 mAbs were conjugated to 40 nm gold nanoparticles (DCN, Carlsbad, CA) through passive adsorption. 1CV18 was excluded from testing as a detection reagent, as the mAb failed to conjugate successfully to the gold nanoparticles. MAb pairs were evaluated in an 18 × 17 matrix to evaluate all capture and detection reagent combinations. Initial testing parameters for assessing capture/detection pairs was 100 ng/mL (positive) and 0 ng/mL (negative) of recombinant SARS-CoV-2 NP (Thermo Fisher Scientific) in 1X DPBS. An ESE Quant GOLD reader (Dialunox, Stockach, Germany) running LF Studio was used to obtain a quantitative intensity signal (mm*mV) at the test line. The signal intensity minus background values were compiled in a heat map (Supplementary Table 5). The top 10 pairs, which exhibited the highest signal over background, were chosen for further LFI development. The selected pairs underwent further testing at 1 ng/mL, 0.5 ng/mL and 0.25 ng/mL of recombinant SARS-CoV-2 (WA1 strain) NP (Thermo Fisher Scientific) in 1X DPBS. In this subsequent testing, the 1CV7 capture and 1CV14 detection combination (1CV7/1CV14) showed the highest reactivity and lowest background of the top ten pairs. As such, 1CV7/1CV14 was selected as the mAb pair for further LFI prototype development.

Initial optimization of the 1CV7/1CV14 LFI was performed with recombinant SARS-CoV-2 (WA1 strain) NP (Thermo Fisher Scientific) diluted in LFI chase buffer containing 1% F127 surfactant (QED Biosciences. San Diego, CA, USA), and 1X DPBS. These conditions were applied to conjugate pad optimization and led to the addition of the 6613H conjugate pad (Ahlstrom-Munksjö, Helsinki, Finland) to the 1CV7/1CV14 LFI. Recombinant SARS-CoV-2 (WA1 strain) NP (Thermo Fisher Scientific) diluted in the pooled nasal matrix collected from normal healthy donors, provided by InBios International Inc., Seattle, WA was used to mimic clinical specimens during the optimization testing.

A panel of viral preparations were acquired from the Biodefense and Emerging Infections Research Resources Repository (BEI Resources) and analyzed on the 1CV7/1CV14 LFI. These viral preparations included rhinovirus (Catalog number NR-51447), human coronavirus (HCoV) OC43 (Catalog number NR-52725), HCoV 229E (Catalog number NR-52726), HCoV NL63 (Catalog number NR-470), Middle East respiratory syndrome Coronavirus (MERS) (Catalog number NR-50171), influenza A (Catalog number NR-19810), influenza B (Catalog number NR-44023), respiratory syncytial virus (RSV) (Catalog number NR-), and SARS-CoV (Catalog number NR-28526). In brief, 20 μL of lysate from cells infected with the respective virus were added to 130 μL of chase buffer. This solution was added to the 1CV7/1CV14 LFI and results were recorded after 20 minutes. All viruses in the panel were ran in duplicate. Rhinovirus, HcoV 229E, MERS, influenza A, influenza B, RSV, and SARS-CoV-2 were tested with of a 1×10^5^ TCID_50_/mL solution. Due to diluted stock concentrations, HCoV OC43 and HCoV NL63 were tested at 8.9×10^4^ TCID_50_/mL and 1.6×10^4^ TCID_50_/mL, respectively. SARS-CoV was tested at 1×10^5^ pfu/mL.

### LFI testing with live SARS-CoV-2 virus

All experiments containing live SARS-CoV-2 were conducted in a biosafety level 3 laboratory. SARS-CoV-2 variants were obtained from the BEI resources: USA-/WA1/2020, Wuhan-Hu1 equivalent; B.1.1.7 (Alpha); C.37 (Lambda); B.1.617.1 (Kappa); B.1.617.2 (Delta); and BA.1 (Omicron). Clinical specimens in the form of remnant patient nasopharyngeal swabs suspended in 1.5 mL of VTM verified via reverse transcriptase-polymerase chain reaction (RT-PCR) to be either COVID-19 positive or negative were provided by Nevada State Public Health Laboratory, Reno, NV. The Ct values of these specimens were determined through RT-PCR using the CDC influenza SARS-CoV-2 Multiplex assay.(26) Pooled normal human nasal matrix, RT-PCR confirmed negative for SARS-CoV-2, was used for contriving specimens for testing on the LFI and BinaxNOW™ COVID-19 Antigen CARD (Abbott, Chicago, IL). For the prototype LFI testing, a determined amount of live virus, denoted in TCID_50_/swab was diluted with pooled normal human nasal matrix to reach 20 μl of the simulated patient sample. The simulated patient sample was then mixed with 130 μl of the chase buffer, applied to the assay and allowed to resolve. Test results were recorded by two observers after 20 mins, and strips were photographed inside the biological safety cabinet. The BinaxNOW™ test was used as recommended (Quick Reference Instructions). Briefly, six drops of the provided chase buffer were added to the card, followed by inserting the provided nasal swab contrived with the indicated amounts (TCID_50_/swab) of the live virus in 20 μl of the nasal matrix. Results were recorded after 15 min and imaged as above. Both 1CV7/1CV14 and BinaxNOW™ LFIs were evaluated with clinical specimens by taking 20 μl of VTM from about 1.5 mL of the total VTM containing nasopharyngeal swabs and adding them to the chase buffer (1CV7/1CV14 LFIs) or onto the BinaxNOW™ swabs. The visible test lines (T) on the 1CV7/1CV14 LFIs and the BinaxNOW™ cards were recorded as positive (+). The test lines that did not show a visible signal were recorded as negative (-). During testing of live virus not from remnant patient samples, barely visible signals were denoted as (+/-) to denote unsure. The BinaxNOW™ test and the 1CV7/1CV14 LFI images were cropped to focus on the test and control lines and assembled for comparative analysis.

### Sequence confirmation of SARS-CoV-2 in clinical specimens

A fraction of the nucleic acid extracted from the VTM with nasopharyngeal swabs was subjected to SARS-CoV-2 sequencing on the Clear Labs DX platform (Clear Labs Inc., San Carlos, CA) utilizing Oxford Nanopore Technology (ONT) or Illumina platform, as described in our previous work. (27–29) The lineages of SARS-CoV-2 present in the nasopharyngeal swabs were defined by the pangolin 4.0.4; PUSHER-v1.2.133 and the sequences were submitted to the GISAID database. VTM-N1 and VTM-N6 (SARS-CoV-2 negative VTMs) were subjected to the extraction of genomic RNA for the detection of viral signatures through sequencing. The sequencing libraries were prepared using the QIAseq FX Single Cell RNA Library kit (QIAGEN, Hilden, Germany), and the SARS-CoV-2 sequences were enriched using a myBaits kit with coronavirus-specific biotinylated probes (Daicel Arbor Biosciences, Ann Arbor, MI.) as described previously. (27) The libraries were sequenced on NextSeq 2000, generated FASTQ files were analyzed, and the mapping reads were visualized using the SARS-CoV-2 mutations analysis tool of the QIAGEN CLC Genomics Workbench (QIAGEN, Inc., Germantown, MD).

## Results

### Monoclonal Antibody production and reactivity

Hybridoma cell lines producing mAbs reactive with SARS-CoV-2 (WA1 strain) NP were cloned twice each by limiting dilution to ensure monoclonality and stability. Ultimately a library of 18 mAbs was created. Reactivity was verified through indirect ELISA. The IgG subclasses of all eighteen mAbs were determined by ELISA with IgG subclass specific mAbs (Supplementary Table 4). Western blot analysis was used to confirm that all eighteen mAbs were reactive to gamma-irradiated lysed Vero E6 cells infected with SARS-CoV-2 (WA1 strain) (data not shown).

### 1CV7/1CV14 LFI demonstrate no cross-reactivity with tested viruses

The 1CV7/1CV14 LFI was evaluated with a cross-reactivity panel of other respiratory viruses: rhinovirus, HCoV OC43, HCoV 229E, HCoV NL63, Middle East respiratory syndrome Coronavirus (MERS), influenza A, influenza B, respiratory syncytial virus (RSV), and SARS-CoV. No reactivity was observed during testing with the 1CV7/1CV14 prototype (Supplementary Figure 2).

### 1CV7/1CV14 LFI and BinaxNOW™ LFI are reactive with multiple SARS-CoV-2 variants

Evaluation of the 1CV7/1CV14 LFI and BinaxNOW™ LFI was performed in triplicate with live WA1, Alpha, Lambda, Kappa, and Delta SARS-CoV-2 (all classified by the CDC as variants of concern) at 500 TCID_50_ diluted into normal human nasal matrix. WA1 and the variants were consistently detected by both assays (Figure 1). Importantly, there was no background signal observed with the 1CV7/1CV14 LFI and BinaxNOW™ when evaluating the normal human nasal matrix alone as a negative control.

**Figure 1.**
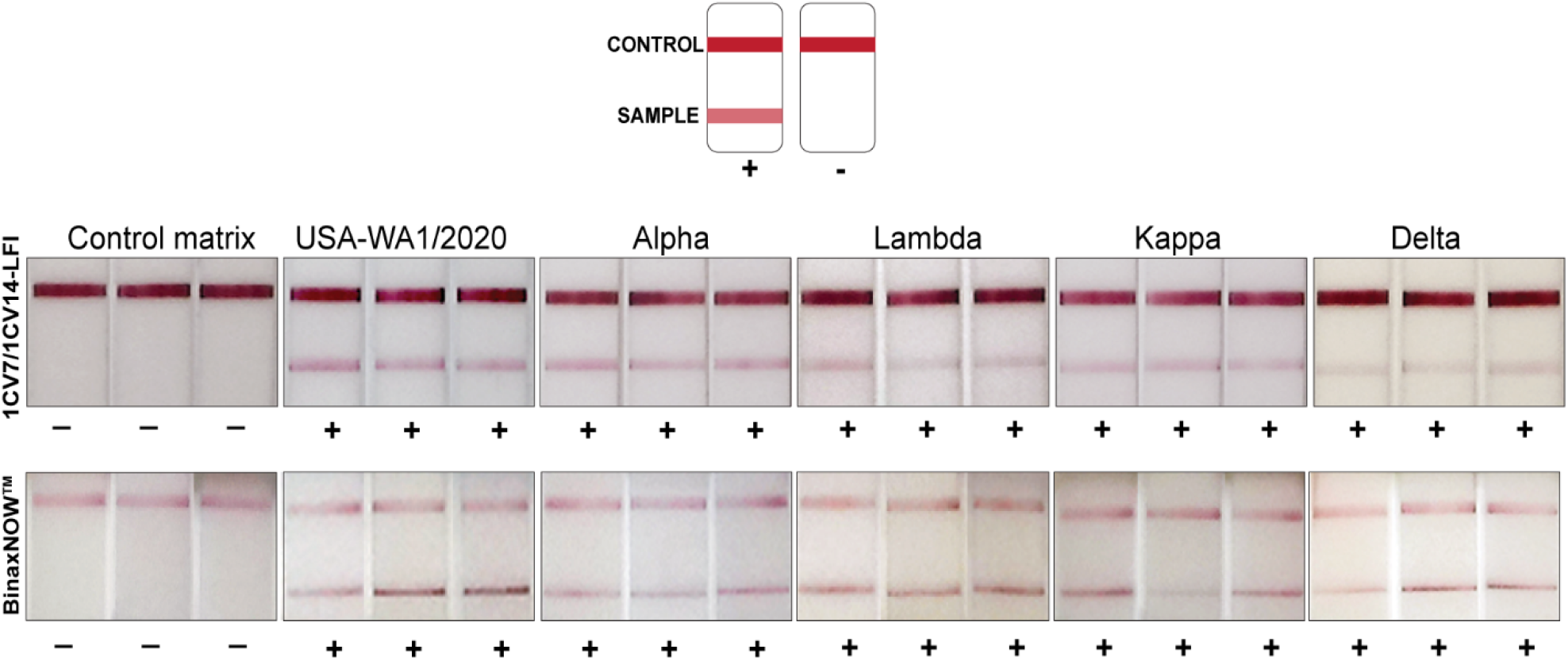
Schematic illustrating positive and negative LFI readings. Additionally, the performance of 1CV7/1CV14 LFI in comparison with BinaxNOW™ for the detection of SARS-CoV-2 variants at 500 TCID_50_/test. A control matrix with known amounts of live SARS-CoV-2 virus was loaded onto the 1CV7/1CV14 LFI as well as the BinaxNOW™ test in triplicates. Signals were recorded through visual inspection of the test lines.

### 1CV7/1CV14 LFI and the BinaxNOW™ LFI have high analytical sensitivity for detection of Delta and Omicron variants

Since Delta and Omicron were the predominant circulating variants at the time of this study, these two variants were used for determining LFI analytical sensitivity. The 1CV7/1CV14 LFI prototype detected the Delta variant at all three concentrations tested (125, 62.5, and 31.25 TCID_50_/swab) (Figure 2A). The BinaxNOW™ tests detected the same concentrations of the Delta variant; however, they were inconsistent in detection in triplicate. The 1CV7/1CV14 LFI also detected the Omicron variant at 15.6 TCID50/swab, which was comparable to the detection limit of the BinaxNOW™ test (Figure 2B). Both LFIs showed weak reactivity at 7.8 TCID_50_/swab with the Omicron variant. Normal human nasal matrix alone was evaluated on both assays in parallel as a negative control, and no background reactivity was observed (Figure 2A).

**Figure 2.**
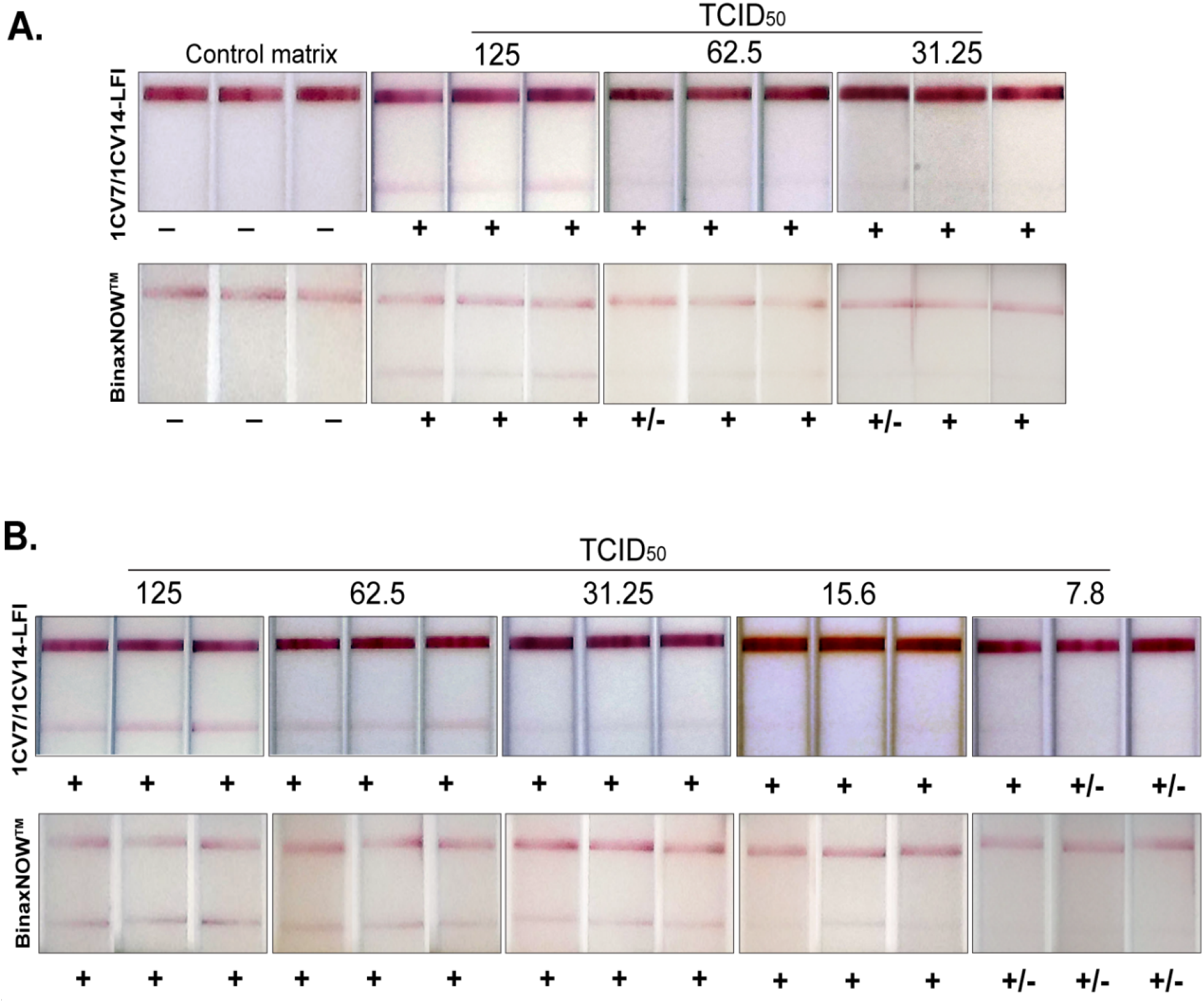
Analytical sensitivities of 1CV7/1CV14 LFI in comparison with BinaxNOW™ for the detection of both Delta (2A) and Omicron BA.1 (2B) variants. A control matrix with known amounts of live SARS-CoV-2 virus of the respective variant was loaded onto the 1CV7/1CV14 LFI as well as the BinaxNOW™ test in triplicate. Signals were recorded through visual inspection of the test lines.

### 1CV7/1CV14 LFI identified more Omicron-positive VTM samples than the BinaxNOW™ LFI

Prior to testing Omicron RT-PCR positive samples, a panel of RT-PCR negative remnant clinical specimens, consisting of nasopharyngeal swabs in VTM collected from symptomatic individuals, were tested on the 1CV7/1CV14 and BinaxNOW™ LFIs (Supplementary Figure 1). 1 RT-PCR negative sample out of the 15 was weakly positive on the 1CV7/1CV14 LFI, while none of the VTMs were positive on the BinaxNOW™. Further investigation into the genomic sequences utilizing an alternative assay (described in the methods section) indicated that the false positive sample (VTM-N6) contained high levels of SARS-CoV-2 RNA, in comparison to another negative sample (VTM-N1) (Supplementary Figure 3).

SARS-CoV-2 Omicron-positive remnants clinical specimens were sequenced and analyzed for lineages through NextClade.(30) Sequences of the clinical isolates were submitted to the GISAID database. A total of 99 samples confirmed to be BA.1 (Omicron), BA.2, BA.2.12.1, BA.4., and BA.5 were assayed on the 1CV7/1CV14 LFI, and results were compared with those of the BinaxNOW™ test. Among 15 BA.1 specimens tested, 10 were positive on the 1CV7/1CV14 LFI and 9 were positive on the BinaxNOW™ LFIs (Figure 3). The 1CV7/1CV14 performed notably better in detecting BA.2; out of the 38 BA.2 samples, 31 samples were detected on the 1CV7/1CV14 LFI compared to 22 on the BinaxNOW™ test (Figure 4A). A subset of 14 samples confirmed to be BA.2.12.1 by RT-PCR (a subvariant of BA.2) resulted in 11 positive 1CV7/1CV14 tests and 9 positives with the BinaxNOW™ (Figure 4B). Both the 1CV7/1CV14 LFI and BinaxNOW™ test were able to detect 3 out of a total of 10 samples confirmed to be Omicron sub-variant BA.4 positive (Figure 5A). When subvariant BA.5 was tested on the two immunoassays, the 1CV7/1CV14 detected 16 samples while the BinaxNOW™ detected 9, out of a total of 21 specimens (Figure 5B). We further compared the test positivity of the clinical specimens with the Ct values determined through RT-PCR using the SARS-CoV-2 and Influenza Multiplex Assay. Across all variants, the 1CV7/1CV14 LFI was able to detect SARS-CoV-2 in clinical specimens with a higher Ct value in comparison to the BinaxNOW™ (Supplementary Table 1, Supplementary Table 2, and Supplementary Table 3). The BinaxNOW™ tests exhibited a higher proportion of negative results.

**Figure 3.**
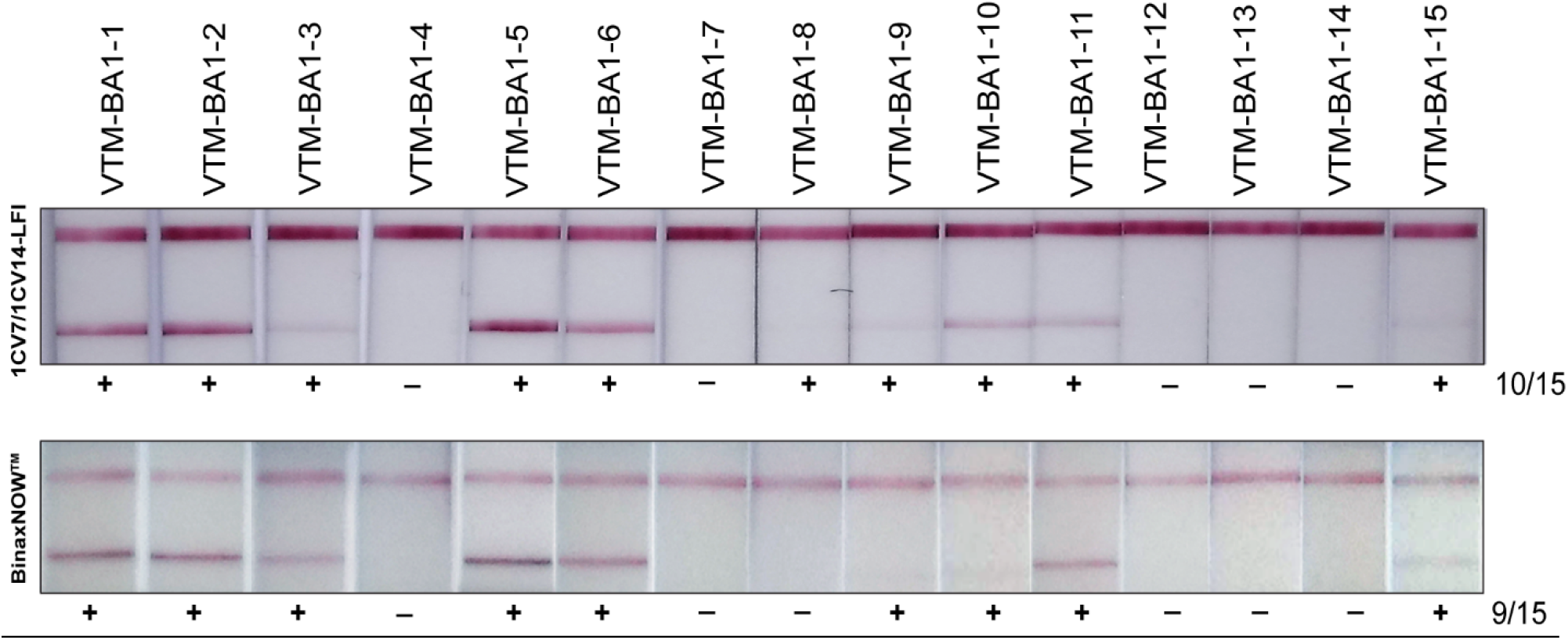
Sensitivities of the 1CV7/1CV14 LFI and BinaxNOW™ in detecting Omicron (BA.1) variant in clinical samples.

**Figure 4.**
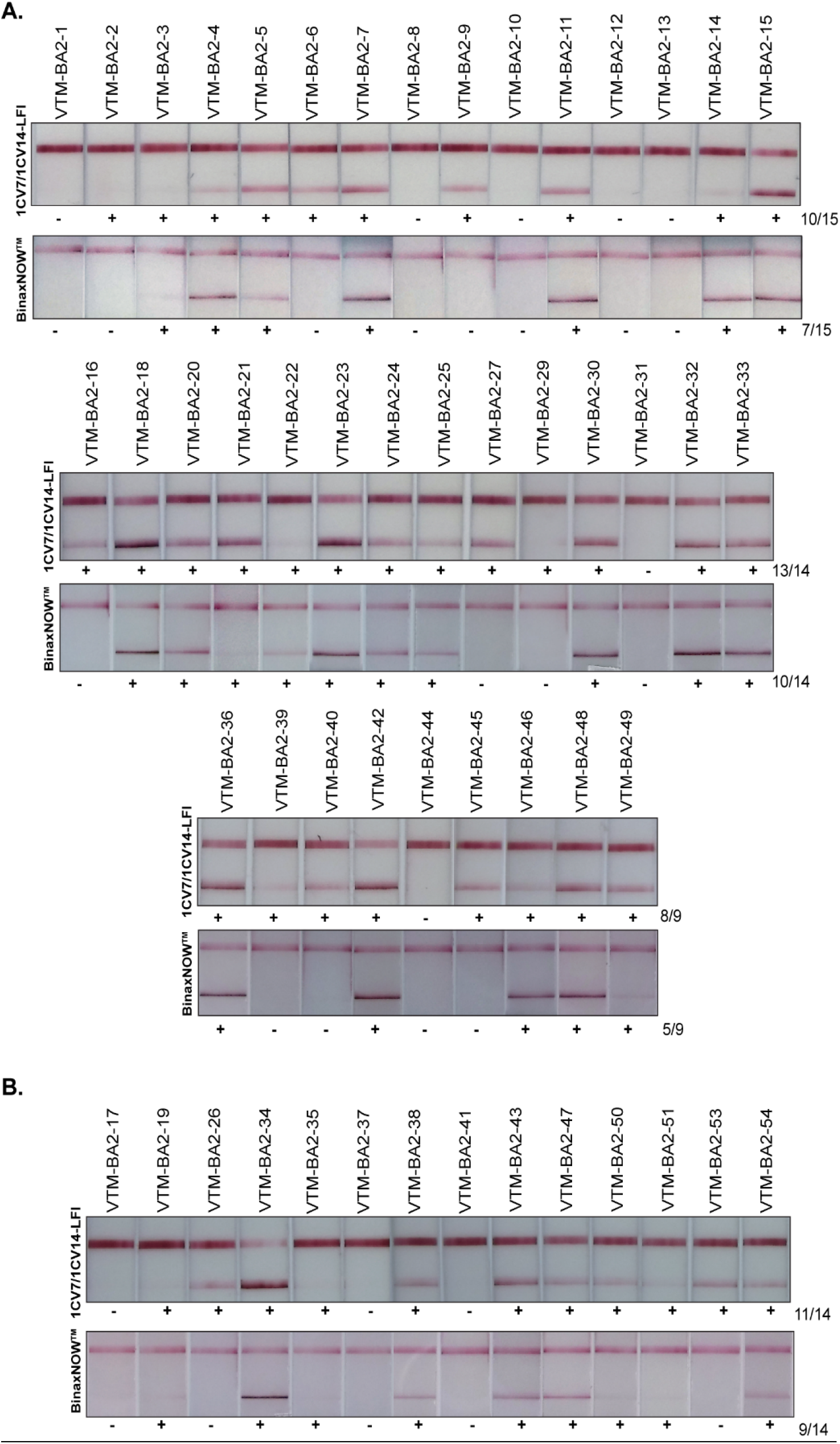
Sensitivities of 1CV7/1CV14 LFIs in comparison with BinaxNOW™ for the detection of Omicron BA.2 (4A) and BA.2.12.1 (4B).

**Figure 5.**
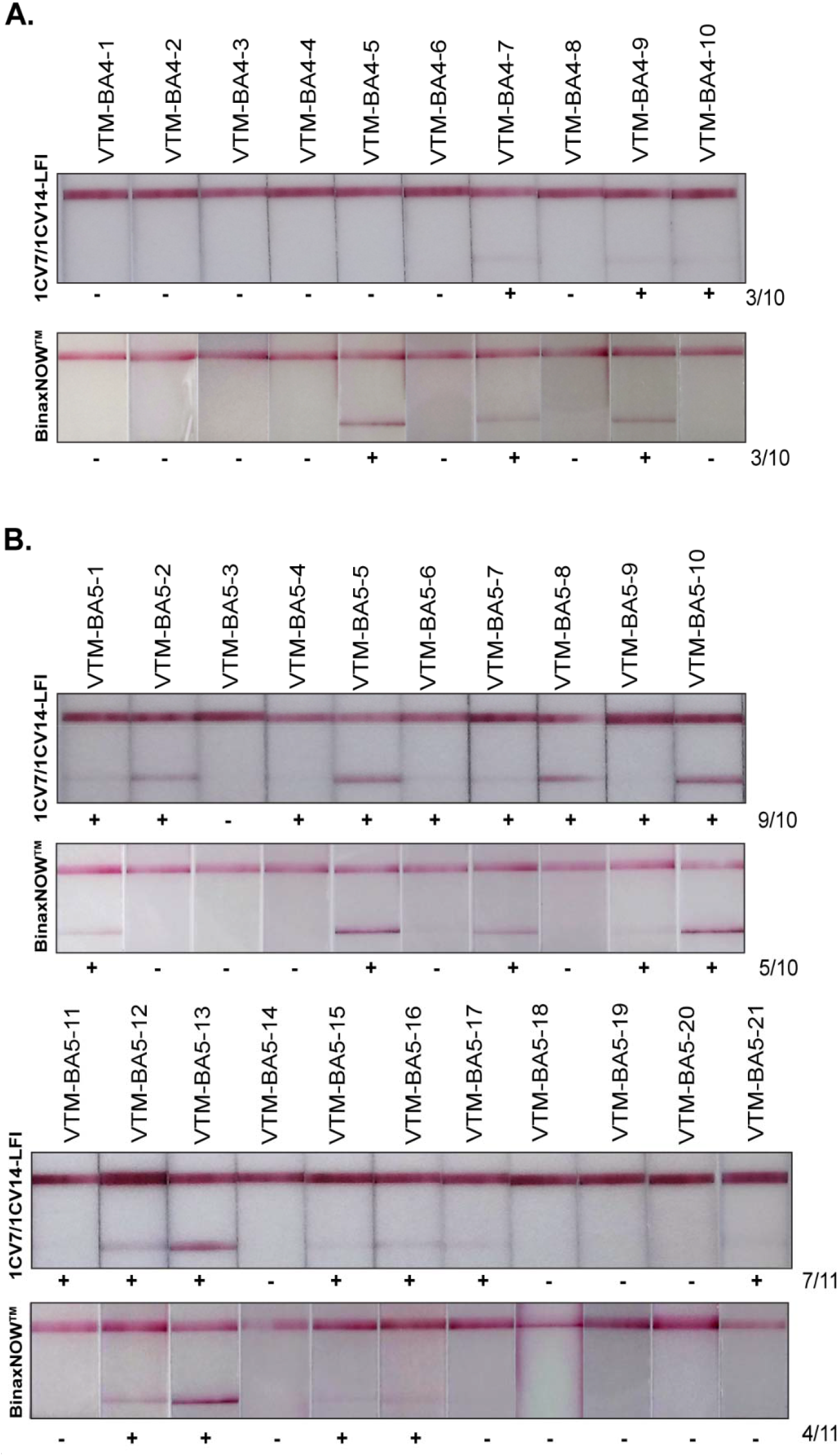
Sensitivities of 1CV7/1CV14 LFIs in comparison with BinaxNOW™ for the detection of Omicron BA.4 (5A) and BA.5 (5B).

## Discussion

The COVID-19 pandemic has underscored the necessity for rapid, accurate, affordable diagnostics that can detect multiple SARS-CoV-2 variants. Leading commercially available rapid antigen tests in the United States are able to detect previous variants of concern.(31) However, currently circulating BA.1, BA.2, BA.2.12.1, BA.4, and BA.5 have shown reduced sensitivity in existing rapid antigen tests.(32) The use of SARS-CoV-2 specific mAbs may prove beneficial for the detection of current and future variants.

In this study, mice were immunized with SARS-CoV-2 (WA1 strain) NP and the library of 18 resulting mAbs was evaluated by LFI. The best performing LFI contained mAbs 1CV7 and 1CV14. Analytical specificity testing of the 1CV7/1CV14 LFI showed no detectable cross-reactivity a small panel of respiratory viruses (Supplementary Figure 2). Notably, the 1CV7/1CV14 LFI did not react with irradiated lysate from SARS-CoV infected cells, indicating our mAbs are recognizing a SARS-CoV-2 NP epitope that is not present on the SARS-CoV NP.

When tested with live virus combined with normal human nasal matrix, at a singular concentration of 500 TCID_50_/test, the 1CV7/1CV14 prototype LFI was able to detect WA1, Alpha, Lambda, Kappa, Delta, and Omicron variants of SARS-CoV-2. Due to Omicron being the most prevalent variant circulating at time of this study, reactivity to this variant was the main focus. The 1CV7/1CV14 LFI achieved an analytical LOD of 15.6 TCID_50_ when testing cultured virus. This was evaluated against the BinaxNOW™ test, which showed a similar LOD (Figure 2B). This is consistent with the two LFIs’ similar test positivity rates with remnant RT-PCR-positive Omicron BA.1 clinical specimens (Figure 2).

When testing remnant, RT-PCR confirmed, Omicron BA.2 or BA.2.12.1 positive patient specimens the 1CV7/1CV14 LFI was able to detect more (42 of 52) of positive specimens than the BinaxNOW™ test (31 of 52). Also consistent with this, the remnant RT-PCR-positive BA.2 or BA.2.12.1 specimens that were detected as positive by the 1CV7/1CV14 LFI had higher RT-PCR Ct values than those that the BinaxNOW™ test was able to detect. The same pattern of greater test positivity rate and detection of specimens with higher Ct values by the 1CV7/1CV14 LFI was also seen with the RT-PCR-positive BA.5 remnant specimens. Additionally, the test lines on the 1CV7/1CV14 LFI appeared more intense, potentially providing easier identification of positive tests, and hence positive specimens. This could be due to mAbs 1CV7 and 1CV14 recognizing more conserved epitopes, or the mAbs simply bind with a higher affinity.

Regarding the detection of BA.4 in clinical samples, both the 1CV7/1CV14 LFI and the BinaxNOW™ test detected 3 of 10 samples (Figure 5A). BA.4 has a unique P151S replacement, in comparison to all other variants.(21) It is possible that the amino acid change from proline to serine results in a slight conformational change in the NP, decreasing the binding ability of the 1CV7, 1CV14, and the BinaxNOW™ antibodies.

Overall, the data presented here demonstrate that the 1CV7/1CV14 LFI is highly inclusive of multiple SARS-CoV-2 strains. Furthermore, the 1CV7/1CV14 LFI appears to be more sensitive in detecting of BA.1, BA.2, BA.2.12.1, and BA.5 in remnant patient specimens when directly compared to the BinaxNOW™ test. The analytical sensitivity of the 1CV7/1CV14 LFI could potentially be futher enhanced by i) adding additional capture or detection mAbs from the existing mAb library (or from other mAb libraries), ii) improving the composition of the chase buffer, and/or iii) the altering the components of the LFI. Furthermore, this study draws attention to the potential importance of developing rapid diagnostic assays with SARS-CoV-2 derived mAbs to hopefully achieve high inclusivity (and hence long-term diagnostic efficacy) across newly evolved SARS-CoV-2 variants of concern.

## Data Availability

The data presented in this manuscript are available from the corresponding author upon reasonable request.

## Contributors

HJH, TU, DH, SGP, JAU, AF, AG, KJP, ARB-M, DER, LG, and SCV contributed in data collection and data analysis. HJH, DH, MAG-H, SCV, and DPA contributed in figure design. HJH, DH, KJP, ARB-M, MAG-H, SCV, and DPA contributed in writing and editing. DPA contributed through study conceptualization, funding acquisition, and supervision.

## Acknowledgments

This work was partially supported by funds from the University of Nevada, Reno’s Research and Innovation Office, DxDiscovery, Inc. internal funds, and through AuCoin Laboratory internal funds.

SARS-CoV-2 isolate USA-WA1/2020 was obtained from the BEI Resources, NIAID, NIH: SARS-Related Coronavirus 2, NR-52281. SARS-CoV-2 variants were also obtained through BEI Resources, NIAID, NIH: SARS-Related Coronavirus 2, Alpha variant, B.1.1.7 (NR-54011), Lambda variant, C37 (NR-55654); Kappa, B.1.617.1 (NR-55486); Delta, B.1.617.2, hCoV-19/USA/MD-HP05285 (NR-55671) and Omicron (Lineage B.1.1.529), hCoV-19/USA/MD-HP20874/2021 (NR-56461). Remnants of clinical specimens sequenced under genomic surveillance study (P20103440) were used for LFI evaluations.

## Figure Legends

**Supplementary figure 1.**
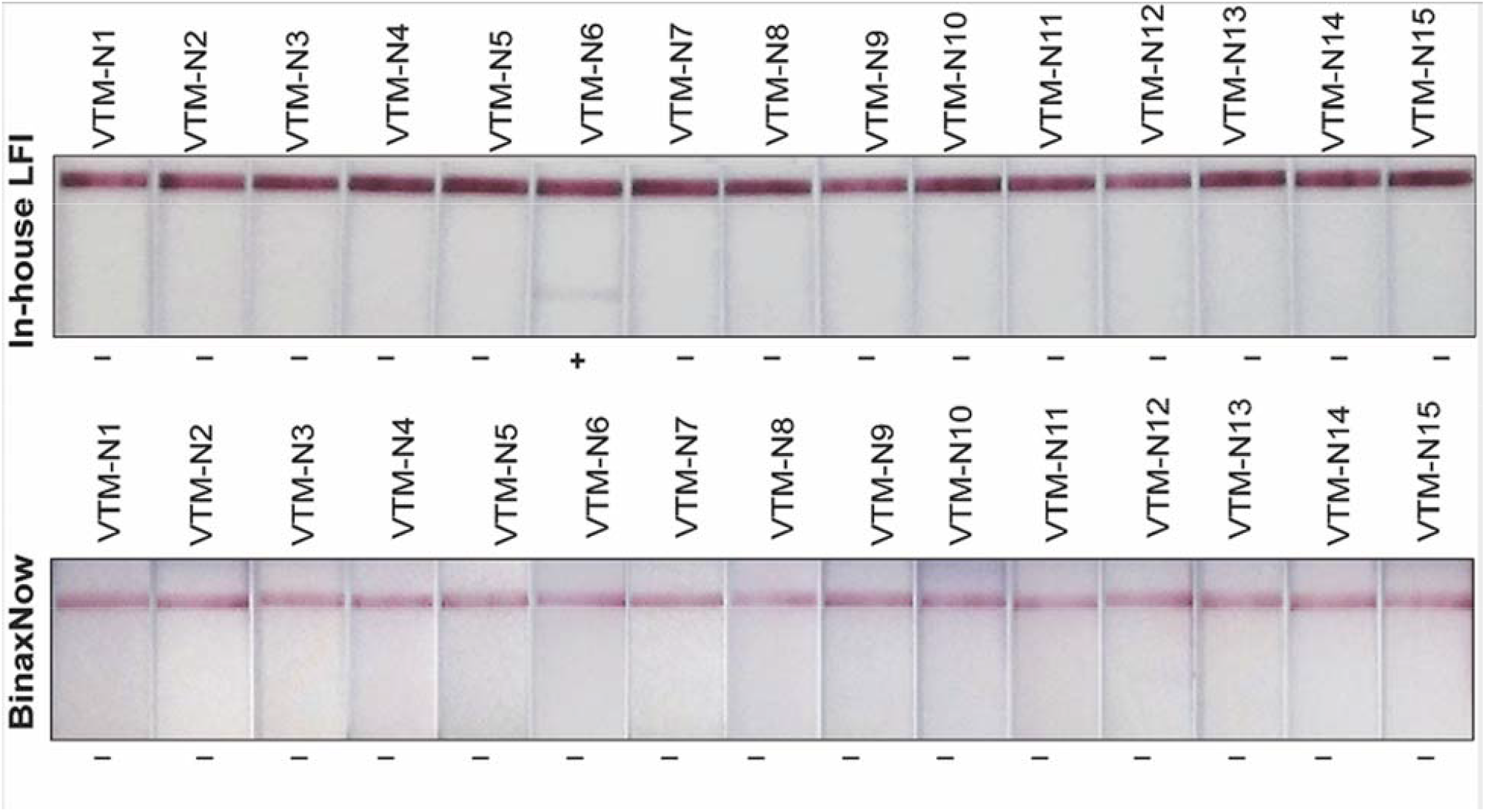
Testing of rt-PCR negative clinical specimens preserved in VTM with the 1CV7/1CV14 LFI and BinaxNOW™ (20 μL sample/test)

**Supplementary figure 2.**
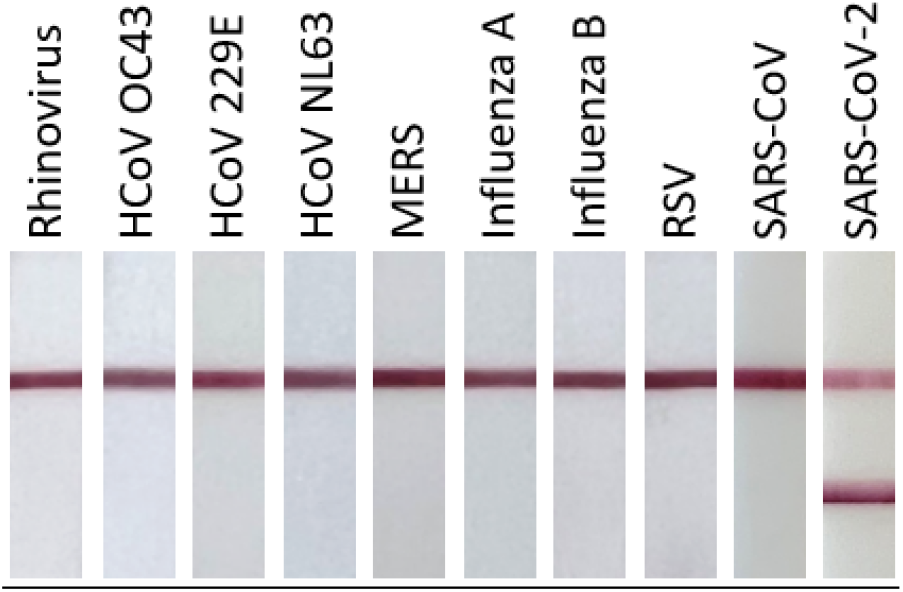
Cross-reactivity testing of 1CV7/1CV14 LFI. Rhinovirus, HCoV 229E, MERS, influenza A, influenza B, RSV, and SARS-CoV-2 concentrations are 1×10^5^ TCID_50_/mL. Due to diluted stock concentrations, HCoV OC43 and HCoV NL63 are at 8.9×10^4^ TCID_50_/mL and 1.6×10^4^ TCID_50_/mL, respectively. SARS-CoV was tested at 1×10^5^ pfu/mL.

**Supplementary figure 3.**
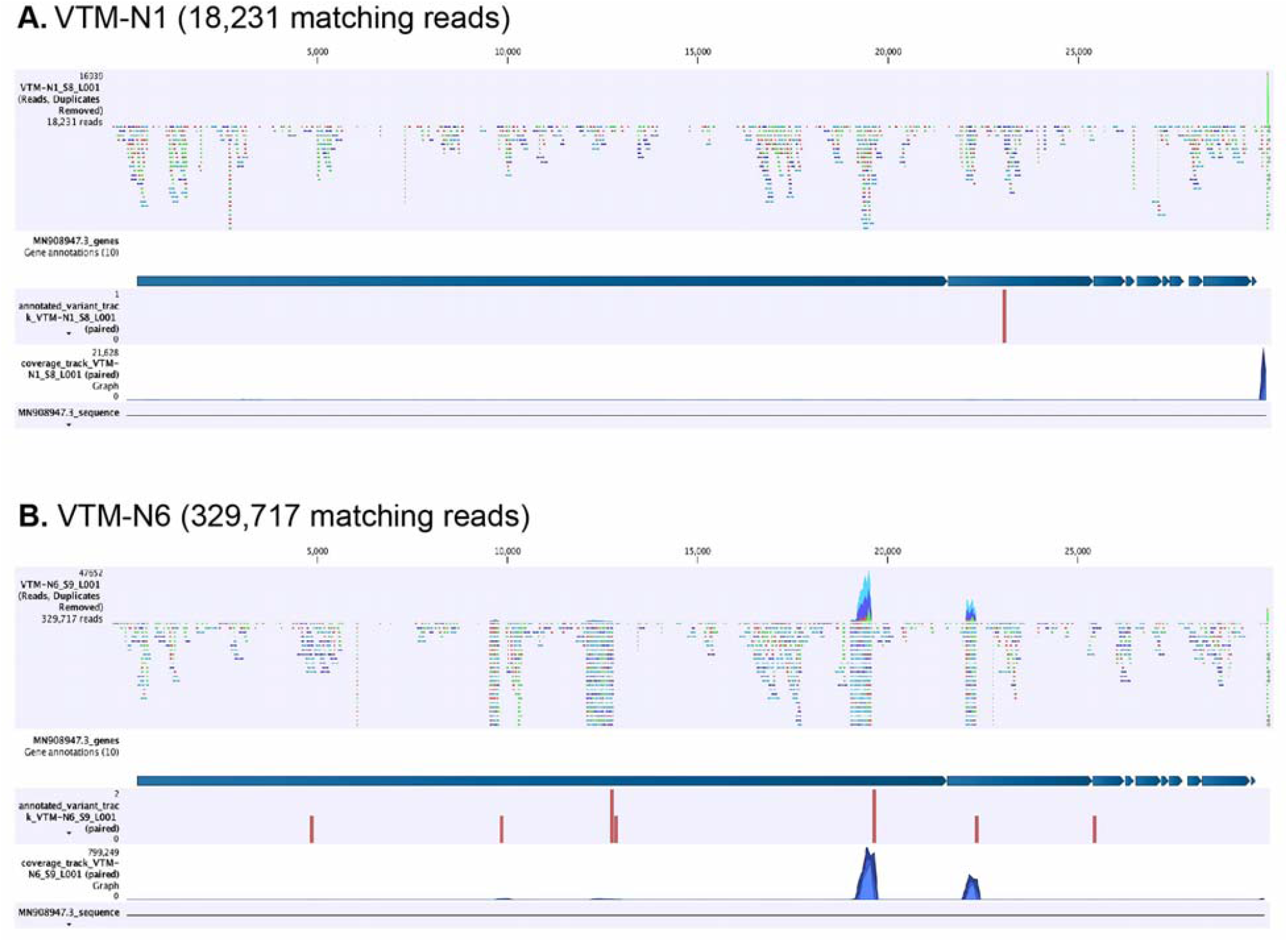
Sequencing of the genomic RNA from the nasopharyngeal swabs of these two specimens showed high levels of the SARS-CoV-2 genome in VTM-N6 as compared to minimal reads in another SARS-CoV-2 negative specimen (VTM-N1).

**Supplementary table 1.** All BA.1 clinical specimens with corresponding Ct values.

| UNR sample IDs | pango_lineage | Ct values | 593 |
| --- | --- | --- | --- |
| VTM-BA1-1 | BA.1.1 | 25.09 |  |
| VTM-BA1-2 | BA.1.1 | 24.32 | 595 |
| VTM-BA1-3 | BA.1.1 | 26.25 |  |
| VTM-BA1-4 | BA.1.1 | 30.06 |  |
| VTM-BA1-5 | BA.1.1 | 20.43 |  |
| VTM-BA1-6 | BA.1.1 | 25.66 |  |
| VTM-BA1-7 | BA.1.1.1 | 35.54 |  |
| VTM-BA1-8 | BA.1 | 32.36 |  |
| VTM-BA1-9 | BA.1.1 | 26.86 |  |
| VTM-BA1-10 | BA.1.15 | 29.56 |  |
| VTM-BA1-11 | BA.1.1 | 26.57 |  |
| VTM-BA1-12 | BA.1 | 27.79 |  |
| VTM-BA1-13 | BA.1.1 | 27.84 |  |
| VTM-BA1-14 | BA.1 | 28.67 |  |
| VTM-BA1-15 | BA.1 | 28.82 |  |

**Supplementary table 2.** All BA.2 and sub-variant BA.2 clinical specimens with corresponding Ct values.

| UNR Sample ID | pango_lineage | Ct values |
| --- | --- | --- |
| VTM-BA2-1 | BA.2 | 23.998 |
| VTM-BA2-2 | BA.2 | 26.182 |
| VTM-BA2-3 | BA.2 | 22.799 |
| VTM-BA2-4 | BA.2 | 15.164 |
| VTM-BA2-5 | BA.2 | 21.678 |
| VTM-BA2-6 | BA.2 | 31.838 |
| VTM-BA2-7 | BA.2.3 | 24.763 |
| VTM-BA2-8 | BA.2 | 30.051 |
| VTM-BA2-9 | BA.2 | 26.558 |
| VTM-BA2-10 | BA.2.3 | 24.626 |
| VTM-BA2-11 | BA.2 | 15.106 |
| VTM-BA2-12 | BA.2 | 19.881 |
| VTM-BA2-13 | BA.2 | 25.742 |
| VTM-BA2-14 | BA.2 | 14.989 |
| VTM-BA2-15 | BA.2 | 20.665 |
| VTM-BA2-16 | BA.2.9 | 23.163 |
| VTM-BA2-18 | BA.2.9 | 15.458 |
| VTM-BA2-20 | BA.2 | 16.75 |
| VTM-BA2-21 | BA.2.3 | 21.354 |
| VTM-BA2-22 | BA.2.3 | 17.214 |
| VTM-BA2-23 | BA.2 | 14.768 |
| VTM-BA2-24 | BA.2.9 | 16.405 |
| VTM-BA2-25 | BA.2 | 16.905 |
| VTM-BA2-27 | BA.2 | 22.68 |
| VTM-BA2-29 | BA.2 | 21.115 |
| VTM-BA2-30 | BA.2.3 | 14.832 |
| VTM-BA2-31 | BA.2 | 28.222 |
| VTM-BA2-32 | BA.2 | 15.229 |
| VTM-BA2-33 | BA.2 | 15.371 |
| VTM-BA2-36 | BA.2 | 15.158 |
| VTM-BA2-39 | BA.2.3 | 26.823 |
| VTM-BA2-40 | BA.2 | 20.231 |
| VTM-BA2-42 | BA.2 | 14.301 |
| VTM-BA2-44 | BA.2 | 22.529 |
| VTM-BA2-45 | BA.2 | 26.262 |
| **VTM-BA2-46** | **BA.2.10** | **17.963** |
| **VTM-BA2-48** | **BA.2** | **17.177** |
| **VTM-BA2-49** | **BA.3** | **21.122** |
| **VTM-BA2-17** | **BA.2.12.1** | **25.148** |
| **VTM-BA2-19** | **BA.2.12.1** | **19.917** |
| **VTM-BA2-26** | **BA.2.12.1** | **20.486** |
| **VTM-BA2-34** | **BA.2.12.1** | **14.365** |
| **VTM-BA2-35** | **BA.2.12.1** | **17.353** |
| **VTM-BA2-37** | **BA.2.12.1** | **26.699** |
| **VTM-BA2-38** | **BA.2.12.1** | **18.873** |
| **VTM-BA2-41** | **BA.2.12.1** | **27.925** |
| **VTM-BA2-43** | **BA.2.12.1** | **18.31** |
| **VTM-BA2-47** | **BA.2.12.1** | **16.462** |
| **VTM-BA2-50** | **BA.2.12.1** | **16.888** |
| **VTM-BA2-51** | **BA.2.12.1** | **19.563** |
| **VTM-BA2-53** | **BA.2.12.1** | **17.416** |
| **VTM-BA2-54** | **BA.2.12.1** | **15.185** |

**Supplementary table 3.**
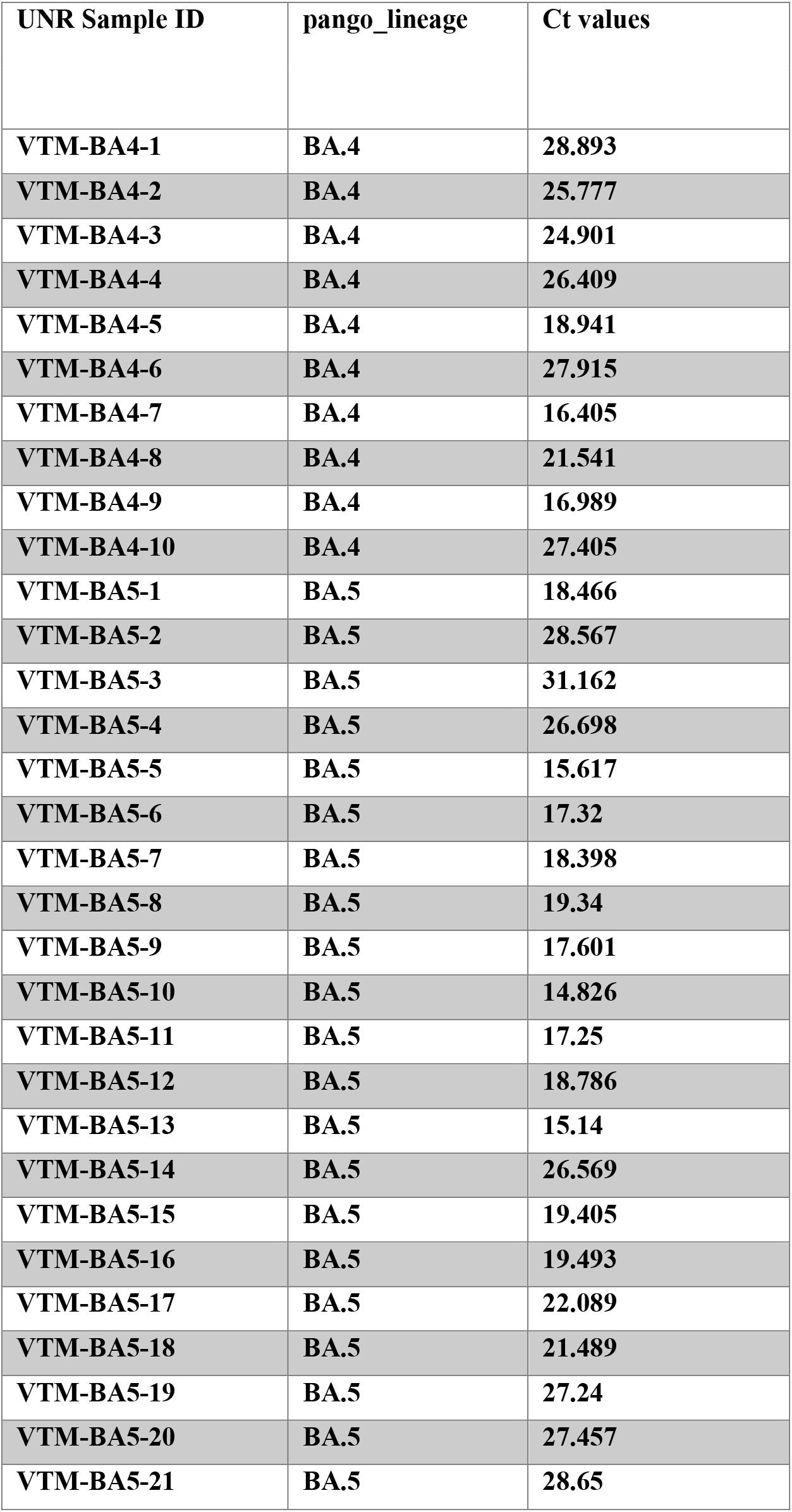
All BA.4 and BA.5 clinical specimens with corresponding Ct values.

| UNR Sample ID | pango_lineage | Ct values |
| --- | --- | --- |
| VTM-BA4-1 | BA.4 | 28.893 |
| VTM-BA4-2 | BA.4 | 25.777 |
| VTM-BA4-3 | BA.4 | 24.901 |
| VTM-BA4-4 | BA.4 | 26.409 |
| VTM-BA4-5 | BA.4 | 18.941 |
| VTM-BA4-6 | BA.4 | 27.915 |
| VTM-BA4-7 | BA.4 | 16.405 |
| VTM-BA4-8 | BA.4 | 21.541 |
| VTM-BA4-9 | BA.4 | 16.989 |
| VTM-BA4-10 | BA.4 | 27.405 |
| VTM-BA5-1 | BA.5 | 18.466 |
| VTM-BA5-2 | BA.5 | 28.567 |
| VTM-BA5-3 | BA.5 | 31.162 |
| VTM-BA5-4 | BA.5 | 26.698 |
| VTM-BA5-5 | BA.5 | 15.617 |
| VTM-BA5-6 | BA.5 | 17.32 |
| VTM-BA5-7 | BA.5 | 18.398 |
| VTM-BA5-8 | BA.5 | 19.34 |
| VTM-BA5-9 | BA.5 | 17.601 |
| VTM-BA5-10 | BA.5 | 14.826 |
| VTM-BA5-11 | BA.5 | 17.25 |
| VTM-BA5-12 | BA.5 | 18.786 |
| VTM-BA5-13 | BA.5 | 15.14 |
| VTM-BA5-14 | BA.5 | 26.569 |
| VTM-BA5-15 | BA.5 | 19.405 |
| VTM-BA5-16 | BA.5 | 19.493 |
| VTM-BA5-17 | BA.5 | 22.089 |
| VTM-BA5-18 | BA.5 | 21.489 |
| VTM-BA5-19 | BA.5 | 27.24 |
| VTM-BA5-20 | BA.5 | 27.457 |
| VTM-BA5-21 | BA.5 | 28.65 |

**Supplementary table 4.**
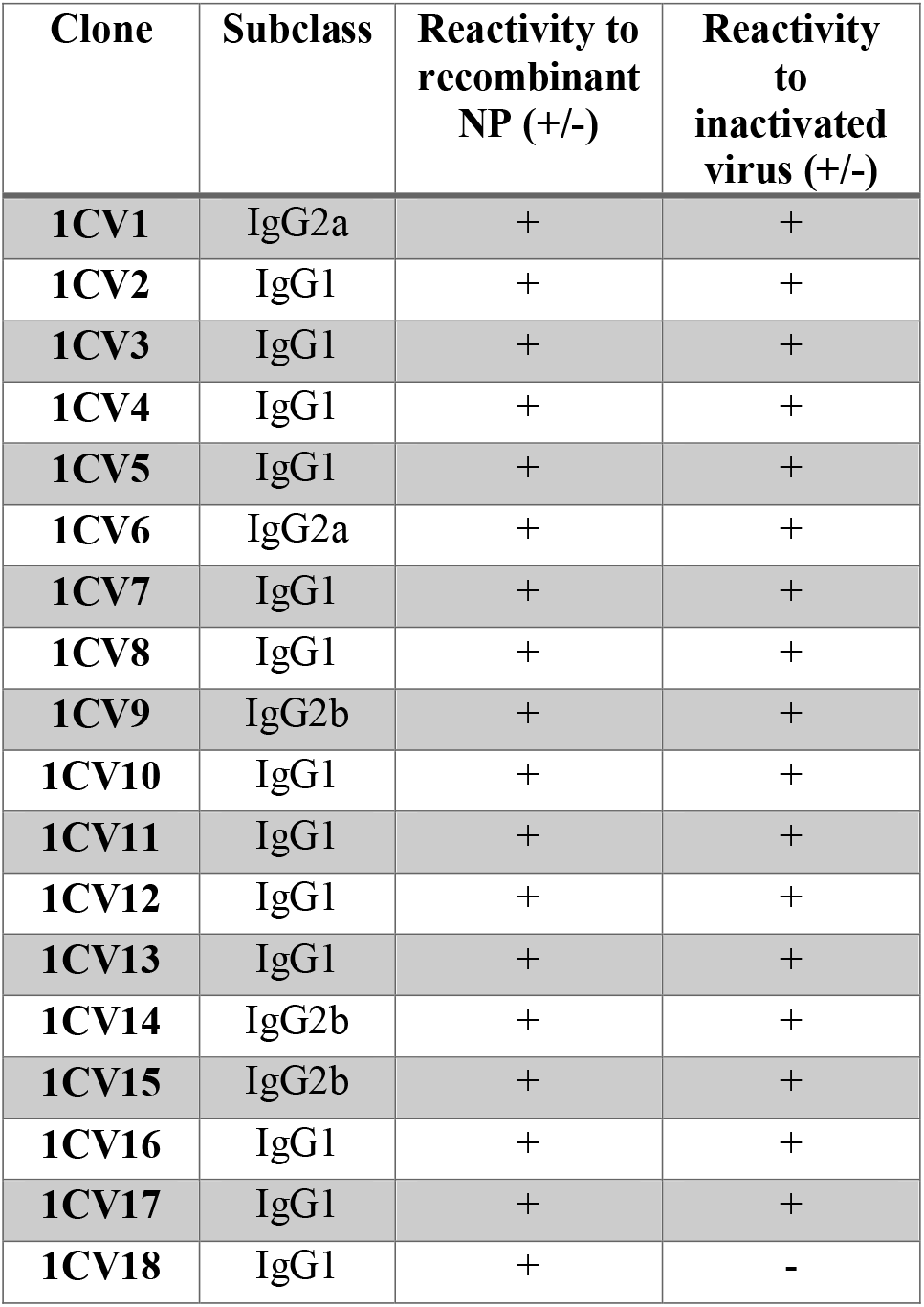
All mAbs generated for mAb library. Subclass is indicated as well as reactivity toward both recombinant NP and viral lysate: “+” indicating reactive and “-” indicating non-reactive.

**Supplementary table 5.**
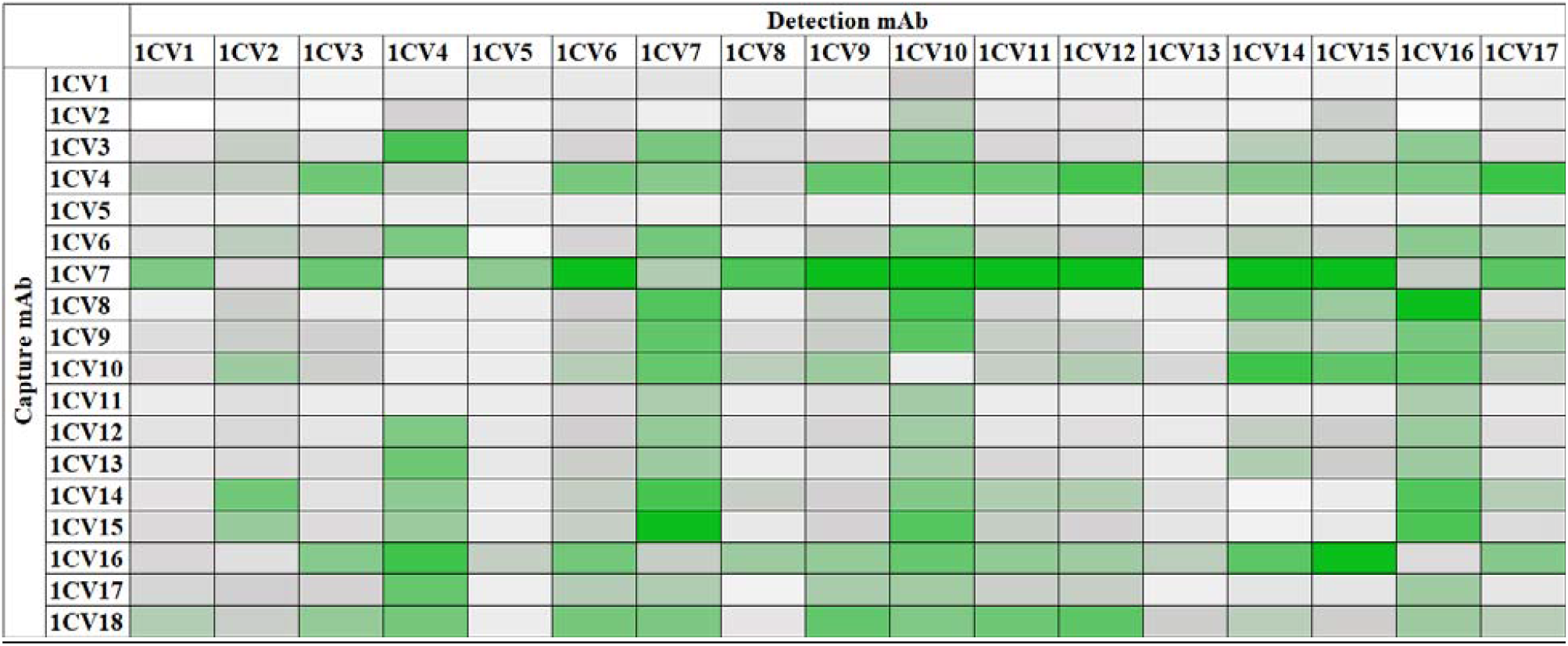
All by all testing of capture/detection pairs. Darker green indicates a higher difference between signal and background.

