## Supplementary figures and images for "Comparison of a prototype SARS-CoV-2 lateral flow immunoassay with the BinaxNOW™ COVID-19 Antigen CARD"

### Supplemental Figure 1

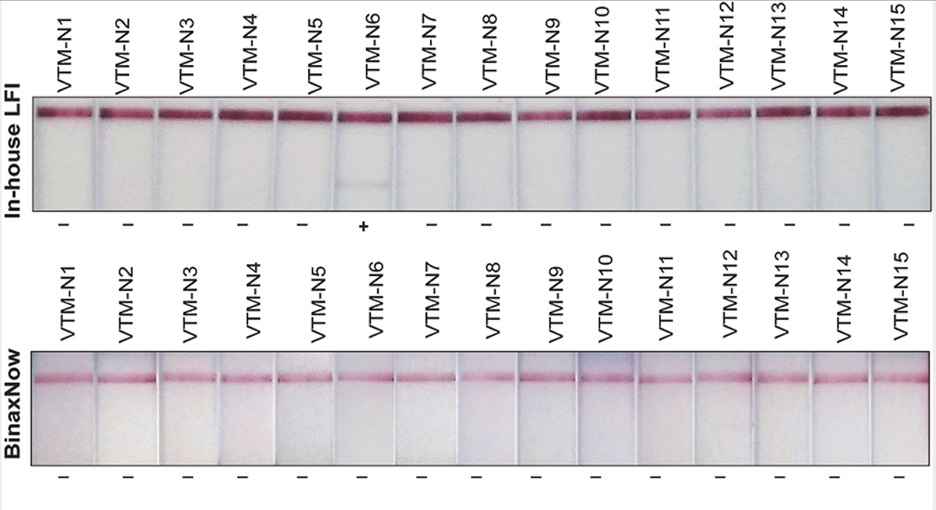

### Supplemental Figure 2

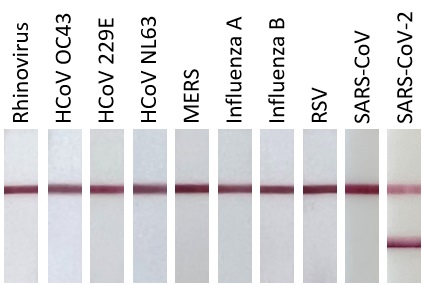

### Supplemental Figure 3

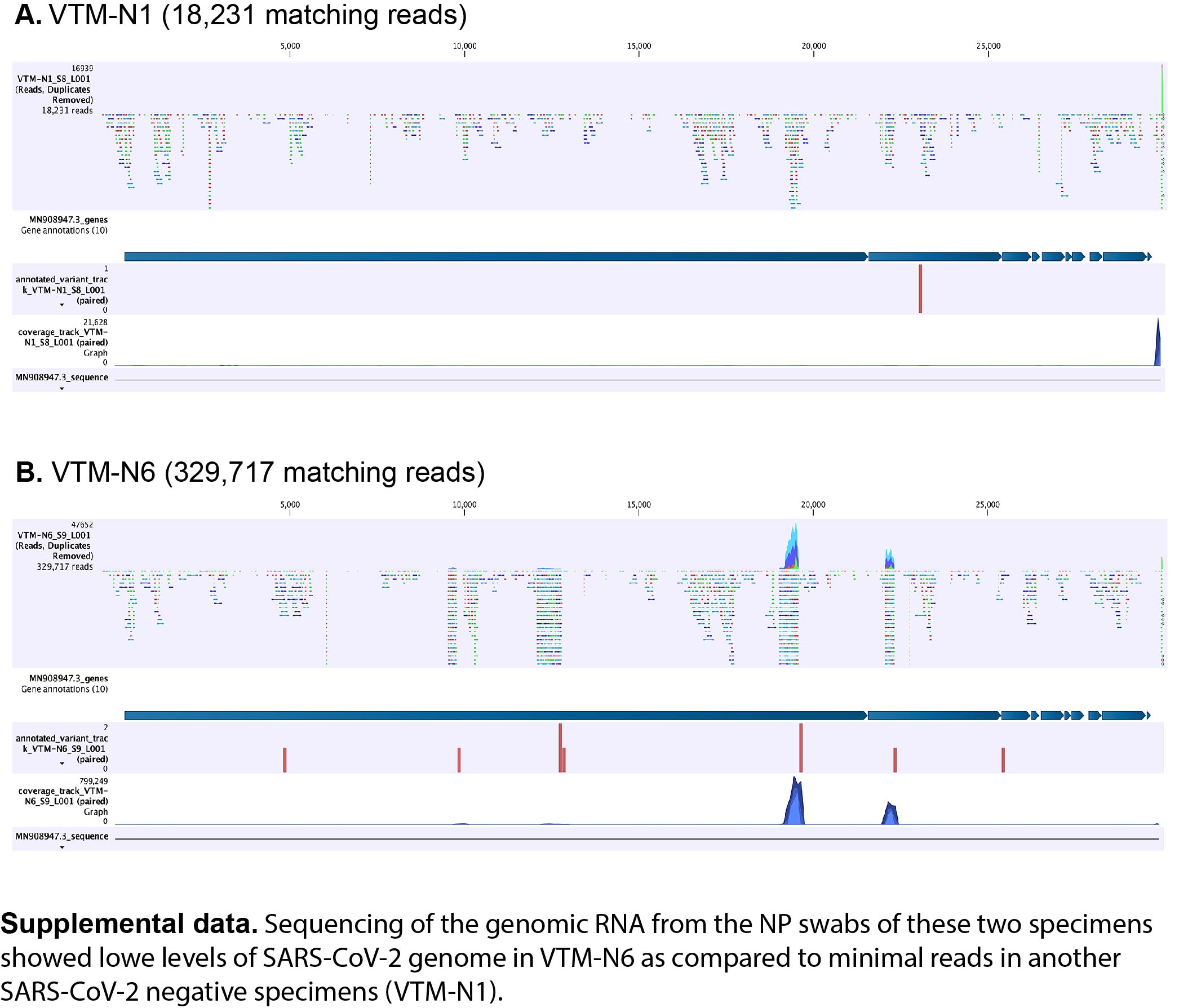
